# Association of Regional Disparity and Socioeconomic Determinants with Acute Malnutrition among Children Aged 6-59 Months in Public Hospitals of Karachi, Pakistan

**DOI:** 10.1101/2025.08.01.25332701

**Authors:** Aisha Gul, Syeda Tabeena Ali, Muhammad Kazim Jafri, Sameera Ali Rizvi

## Abstract

**Objective:** This study aimed to examine the association of regional disparity and socioeconomic determinants with acute malnutrition among children aged 6-59 months attending selected public hospitals in Karachi.

**Methods:** A hospital-based unmatched case-control study was conducted on 394 participants with 197 cases and 197 controls. After developing a self-structured questionnaire based on maternal and child components, wealth quintiles were made using principal component analysis. A univariate and then a binary logistic regression was applied.

**Results:** The odds of acute malnutrition were 1.2 times higher in male children. The variables significantly associated with childhood acute malnutrition were regional disparity [AOR=2.3, 95%, C. I (1.4-3.8), p-value <0.01], mother’s illiteracy [AOR = 3.6, 95% C. I (1.9-7), p-value <0.001], mother’s primary education [AOR =1.1, 95% C. I (0.5-2.2), p-value <0.01], father’s illiteracy [AOR= 2.4, 95% C. I (0.8-4.4), p-value 0.03], father’s primary education [AOR =1.1, 95% C. I (0.4-1.9), p-value <0.01], poorest households [AOR= 2.2, 95% C. I (1.07-4.7), p-value <0.01], and child’s age [AOR =0.945, 95% C. I (0.92-0.96), p-value <0.01].

**Conclusion:** Regional disparity was found to be significantly associated with acute malnutrition among children along with household wealth status, mother’s illiteracy, and father’s illiteracy. Hence there is a need to direct the focus of policy makers to work on these factors to eradicate acute malnutrition among children.

## Introduction

Malnutrition encompasses both overabundant and or insufficient nutritional intake, which results in an imbalance between nutrient intake and its utilization [1].

Acute malnutrition occurs due to inadequate nutritional intake for under three months. It manifests as moderate acute malnutrition (MAM) when weight-for-height is between −2 and −3 SD or mid-upper arm circumference (MUAC) is <12.5 cm (125 mm). Severe acute malnutrition (SAM) is diagnosed when weight-for-height is < - 3 SD or MUAC is <11 cm (115 mm) [2]. In 2019, 30 million children worldwide suffered severe malnutrition, primarily in countries facing crises, pandemics, climate change, and rising living costs. Acute malnutrition affected 17% of Pakistani children under five, with Sindh showing a higher prevalence of 23.3 % in 2019. According to the report generated by the National Nutritional Survey in 2018, acute malnutrition, in the form of wasting was 18.4% prevalent in boys [3]. A country’s economic progress is connected to regional disparities, which include geographical, social, and economic inequality [4]. In Pakistan, rural areas have higher wasting rates (18.6%) than urban areas. However, a recent MICS-based study in Multan found a higher prevalence in urban areas [5]. Acute malnutrition has been a public health concern for developing countries like Pakistan, where scarcity of resources makes things worse, especially in terms of nutrition. Acute malnutrition in children (low weight-for-height Z-score < −2 SD or MUAC <125 mm) is influenced by illiteracy, large families, poor breastfeeding, poverty, early weaning, nuclear families, lack of immunization, teenage pregnancies, overcrowding, and inadequate child healthcare [6].

A study in Africa (2010-2018) using demographic health survey data from 51 low-middle-income countries, including Pakistan, found severe acute malnutrition more prevalent in rural areas, except in Tajikistan and Malawi [7]. While published literature is scarce on regional disparities and acute malnutrition in, Pakistan. A study conducted in Faisalabad, Pakistan highlighted that rural areas had a 33% prevalence of child malnutrition, pointing towards regional disparity as a contributor to childhood malnutrition [6]. Acute malnutrition threatens children’s developmental and cognitive abilities, burdening national health and economy [8]. The Sindh Bureau of Statistics found children of uneducated mothers are more prone to wasting [9]. While linked to regional disparities, research on its socioeconomic determinants remains limited. Parental education and employment, which influence household wealth, contribute to wasting. As per the given facts, acute malnutrition among children is increasing at alarming rates. And since there is a lack of comprehensive research on how regional disparities and socioeconomic determinants contribute to acute malnutrition, this is expected to burden the country’s social and economic development. Child well-being is essential for national development, particularly during the 6-59 months for cognitive growth. In Pakistan, children under five make up 15% of the population but account for 50% of child mortality [10]. Acute malnutrition is a major contributor, potentially causing nearly half of these deaths if untreated [11]. While maternal sociodemographic factors have been studied before, there is limited research on how regional disparities affect malnutrition in Karachi. This study aimed to fill this gap by exploring the association between regional disparities, socioeconomic status, and acute malnutrition in children aged 6-59 months. By analyzing wealth quintiles and comparing children with normal nutritional status to those affected by malnutrition, the research will provide new insights. The findings will inform policymakers, helping to shape or revise policies to improve child nutrition and reduce malnutrition-related mortality. The study will help to investigate and contribute to the body of literature about the association of regional disparity and socioeconomic determinants with acute malnutrition among children aged 6-59 months, attending selected public hospitals in Karachi, Pakistan.

## Methods

This hospital-based, unmatched case-control study was conducted at Civil Hospital Karachi (CHK) and Public Health School Karachi (PHS) from September 2023 to May 2024. These hospitals were chosen for their high pediatric outpatient volume and diverse socioeconomic patient base, facilitating the investigation of factors related to acute malnutrition. Participants included children aged 6–59 months, residing in Karachi for at least six months. Cases had acute malnutrition (WFH z-score < −2 SD), and controls had normal nutritional status (−2 SD to +2 SD). Nutritional status was assessed using weight-for-height Z-scores, a gold standard method for clinical malnutrition [12]. Weight and height were measured following standard protocols.

Exclusion criteria included chronic illnesses, preterm birth, congenital abnormalities, and refusal of consent. Ethical approvals were obtained from SZABIST University’s Ref. No. IERB(1 8)/S ZARIST-KHI(PH)I 22104145 1240067, Civil Hospital (MS/CHK/17654), and Public Health School (PHS/1318).

The sample size was calculated using EPI-Info version 7, requiring 358 participants, adjusted to 394 for non-response. Based on pediatric OPD visits, 88.5% (350 participants: 175 cases, 175 controls) were allocated to CHK, and 11.5% (44 participants: 22 cases, 22 controls) to PHS. The questionnaire was pre-tested on 10% of the sample (36 participants) at CHK, ensuring clarity and validity, with necessary modifications.

The dependent variable was acute malnutrition based on WFH Z-scores. Independent variables included socio-demographic and socioeconomic factors such as household type, asset ownership, health access, and water source. Descriptive analysis categorized WFH Z-scores into normal nutrition and acute malnutrition.

Data were analyzed using SPSS Version 22. Descriptive statistics were computed for all variables. The inferential analysis included independent t-tests for categorical-continuous variables. Pearson’s Chi-square and Fisher’s exact test assessed relationships between categorical variables, with p < 0.05 considered significant.

Binary logistic regression identified factors associated with acute malnutrition, adjusting for other variables. Variables with p < 0.05 in univariate analysis were included.

### Ethical Considerations

Written informed consent was obtained from mothers, with the study’s purpose, risks, and benefits explained in understandable language. Confidentiality was ensured, and participants could withdraw anytime without consequences. Selection was non-discriminatory, and children’s well-being was monitored throughout. Ethical approval was granted by SZABIST University. No. IERB (1 8)/S ZARIST-KHI(PH)I 22104145 1240067, and the data was collected from Civil Hospital and Public Health School, Karachi. Participation was voluntary, with no harm or loss of benefits for refusal. Data was securely stored and used solely for research.

## RESULTS

Among 394 participants (1:1 case to controls), 58.4% were male, and 41.6% were female. Cases included 60.9% males and 39.1% females. Ages ranged from 6–54 months (mean 25 ± 12.59). Most participants (65.5%) lived in the main city, while 34.5% were from slums. Among cases, 43.1% were from slums.

Multivariate analysis showed significant associations with slum residence [AOR = 2.3, CI (1.4–3.8), p = 0.001], maternal illiteracy [AOR = 3.6, CI (1.9–7), p = 0.001], paternal illiteracy [AOR = 2.4, CI (0.8–4.4), p = 0.03], and poverty [AOR = 2.2, CI (1.07–4.7), p = 0.01]. An inverse relationship was observed between child age and malnutrition; older children had lower odds of malnutrition [AOR = 0.94, CI (0.92–0.96), p = 0.001].

The analysis highlighted strong associations between acute malnutrition and factors like regional disparity, parental education, wealth status, and child age. Slum residence and poverty were key contributors, with slum children showing 2.3 times higher odds of malnutrition. Children of non-educated mothers were 3.6 times more likely to be malnourished, and those from poorer households had 2.2 times higher odds compared to richer households.

**Table 1:** Descriptive statistics of maternal and child characteristics among cases and controls.

| Variable | Cases (n=197) | Controls (n=197) |
| --- | --- | --- |
| **Maternal Demographics** |  |  |
| **Age of Mother** |  |  |
| Less than 24 | 27 (13.7%) | 38 (19.3%) |
| 24 and above | 170 (86.3%) | 159 (80.7%) |
| **Residential Area** |  |  |
| Slum | 85 (43.1%) | 51 (25.9%) |
| Main City | 112 (56.9%) | 146 (74.1%) |
| **House Type** |  |  |
| Cemented | 166 (84.3%) | 185 (93.9%) |
| Non-Cemented | 31 (15.7%) | 12 (6.1%) |
| **Number of Rooms** |  |  |
| ≤3 | 166 (84.3%) | 150 (76.2%) |
| >3 | 31 (15.7%) | 47 (23.8%) |
| **Family Size** |  |  |
| <5 | 63 (32%) | 50 (25.4%) |
| ≥5 | 134 (68%) | 147 (74.6%) |
| **Toilet Type** |  |  |
| Cemented | 166 (84.3%) | 185 (93.9%) |
| Non-Cemented | 31 (15.7%) | 12 (6.1%) |
| **Parental Education** |  |  |
| **Mother's Education** |  |  |
| None | 141 (71.6%) | 86 (43.7%) |
| Primary | 35 (17.7%) | 64 (32.5%) |
| Secondary+ | 21 (10.7%) | 47 (23.8%) |
| **Father's Education** |  |  |
| None | 49 (24.9%) | 16 (8.1%) |
| Primary | 44 (22.3%) | 50 (25.4%) |
| Secondary+ | 104 (52.8%) | 131 (66.5%) |
| **Parental Employment & Income** |  |  |
| **Employment Status (Mother)** |  |  |
| Employed | 26 (13.2%) | 21 (10.7%) |
| Unemployed | 171 (86.8%) | 176 (89.3%) |
| **Employment Status (Father)** |  |  |
| Employed | 188 (95.4%) | 193 (98%) |
| Unemployed | 9 (4.6%) | 4 (2%) |
| **Income** |  |  |
| ≤20,000 | 147 (74.6%) | 56 (38.6%) |
| 20,001–50,000 | 46 (23.4%) | 111 (56.3%) |
| >50,000 | 4 (2%) | 10 (5.1%) |
| **Wealth Quintiles** |  |  |
| Poorest | 55 (27.9%) | 31 (15.7%) |
| Lower Middle | 49 (24.9%) | 28 (14.2%) |
| Middle | 34 (17.3%) | 45 (22.8%) |
| Upper Middle | 31 (15.7%) | 47 (23.9%) |
| Rich | 28 (14.2%) | 46 (23.4%) |
| **Marriage & Index Pregnancy** |  |  |
| **Marital Status** |  |  |
| Married | 189 (95.9%) | 193 (97.9%) |
| Separated/Divorced/Widowed | 8 (4.1%) | 4 (2.1%) |
| **Age at Index Pregnancy** |  |  |
| ≤24 | 28 (14.2%) | 39 (19.8%) |
| ≥25 | 169 (85.8%) | 158 (80.2%) |
| **Number of Under-5 Children** |  |  |
| 1 | 110 (55.8%) | 118 (59.9%) |
| 2 | 81 (41.1%) | 78 (39.6%) |
| 3 | 6 (3%) | 1 (0.5%) |
| **Sanitation & Hygiene** |  |  |
| **Drinking Water Source** |  |  |
| Tap Water | 138 (70.1%) | 145 (73.6%) |
| Filter/Dispenser | 55 (27.9%) | 51 (25.9%) |
| Others | 4 (2%) | 1 (0.5%) |
| **Water Purification** |  |  |
| Yes | 96 (48.7%) | 118 (59.9%) |
| No | 101 (51.3%) | 79 (40.1%) |
| **Handwashing Before Feeding** |  |  |
| Yes | 12 (6.1%) | 193 (98%) |
| No | 185 (93.9%) | 4 (2%) |
| **Toilet Facility** |  |  |
| Shared | 130 (66%) | 117 (59.4%) |
| Separate | 67 (34%) | 80 (40.6%) |
| **Fecal Material Disposal** |  |  |
| Flush | 175 (88.8%) | 189 (95.9%) |
| Pit Latrine | 22 (11.2%) | 8 (4.1%) |
| **Child Nutrition** |  |  |
| **First Diet** |  |  |
| Formula Milk | 94 (47.7%) | 79 (40.1%) |
| Mother's Milk | 103 (52.3%) | 118 (59.9%) |
| **Breastfed Timing** |  |  |
| Within 1 Hour | 65 (33%) | 108 (54.8%) |
| After 1 Hour | 132 (67%) | 89 (45.2%) |
| **Prelacteal** |  |  |
| Yes | 126 (64.5%) | 107 (54.3%) |
| No | 70 (35.5%) | 90 (45.7%) |
| **EBF Duration** |  |  |
| <5 Months | 144 (73.1%) | 15 (7.6%) |
| ≥5 Months | 53 (26.9%) | 182 (92.4%) |
| **Weaning Diet** |  |  |
| Wheat | 95 (48.2%) | 62 (31.5%) |
| Rice | 39 (19.8%) | 50 (25.4%) |
| Semolina | 27 (13.7%) | 54 (27.4%) |
| Fruits | 18 (9.1%) | 12 (6.1%) |
| Dairy Items | 18 (9.1%) | 19 (9.6%) |
| **Child Pro-forma** |  |  |
| **Mode of Birth** |  |  |
| NVD | 93 (47.2%) | 120 (60.9%) |
| LSCS | 104 (52.8%) | 77 (39.1%) |
| **Religion** |  |  |
| Muslim | 170 (86.3%) | 186 (94.4%) |
| Non-Muslim | 27 (13.7%) | 11 (5.6%) |
| **Gender** |  |  |
| Male | 120 (60.9%) | 110 (55.8%) |
| Female | 77 (39.1%) | 87 (44.2%) |
| **Birth Order** |  |  |
| Less than or equal to 3 | 184 (93.4%) | 188 (95.4%) |
| More than 3 | 13 (6.6%) | 9 (4.6%) |
| **Age of Child (Months)** |  |  |
| 6–11 | 8 (4.1%) | 12 (6.1%) |
| 12–23 | 90 (45.7%) | 51 (25.9%) |
| 24–35 | 62 (31.5%) | 52 (26.4%) |
| 36–47 | 24 (12.2%) | 44 (22.3%) |
| 48–59 | 13 (6.6%) | 38 (19.3%) |
| **Birth Weight (kg)** |  |  |
| Mean ± S.D. | 2.8 ± 0.28 | 3.2 ± 0.26 |
| **Current Weight (kg)** |  |  |
| Mean ± S.D. | 8.98 ± 1.5 | 11.7 ± 2.7 |
| **Height (cm)** |  |  |
| Mean ± S.D. | 83.8 ± 8.3 | 84.2 ± 11.23 |
| **Weight-for-Height Z-score** |  |  |
| Mean ± S.D. | -2.66 ± 0.97 | 0.59 ± 1.37 |

**Table 2:** Crude Odds of Acute Malnutrition by Child Variables.

| Variable | Category | Cases (n=197) | Controls (n=197) | COR | 95% CI | p-value |
| --- | --- | --- | --- | --- | --- | --- |
| **Mode of Birth** | NVD | 93 (47.2%) | 120 (60.9%) | 0.89 | 0.59–1.5 | 0.479 |
|  | LSCS* | 104 (52.8%) | 77 (39.1%) | Ref | – | – |
| **Birth Order** | ≤3 | 184 (93.4%) | 188 (95.4%) | 0.6 | 0.28–1.6 | 0.38 |
|  | >3* | 13 (6.6%) | 9 (4.6%) | Ref | – | – |
| **Age of Child** | 6–11 | 8 (4.1%) | 12 (6.1%) | 2.44 | 0.85–6.97 | 0.097 |
|  | 12–23 | 90 (45.7%) | 51 (25.9%) | 6.39 | 2.69–15.1 | <0.001 |
|  | 24–35 | 62 (31.5%) | 52 (26.4%) | 4.36 | 1.80–10.6 | 0.001 |
|  | 36–47 | 24 (12.2%) | 44 (22.3%) | 2.00 | 0.78–5.09 | 0.142 |
|  | 48–59* | 13 (6.6%) | 38 (19.3%) | 1.00 | – | – |
| **Gender** | Male | 120 (60.9%) | 110 (55.8%) | 1.2 | 0.8–1.8 | 0.51 |
|  | Female* | 77 (39.1%) | 87 (44.2%) | Ref | – | – |
| **Religion** | Muslim | 170 (86.3%) | 186 (94.4%) | 1 | 0.3–2.7 | 0.9 |
|  | Non-Muslim* | 27 (13.7%) | 11 (5.6%) | Ref | – | – |

\*Reference category

**Table 3:** Multivariate Logistic Regression — Association of Sociodemographic Variables with Acute Malnutrition.

| Variable | Cases (n=197) | Controls (n=197) | AOR | 95% CI | p-value |
| --- | --- | --- | --- | --- | --- |
| **Residential Area** |  |  |  |  |  |
| Slum | 85 (43.1%) | 51 (25.9%) | 2.3 | 1.4–3.8 | 0.001 |
| Main City* | 112 (56.9%) | 146 (74.1%) | Ref | – | – |
| **Mother's Education** |  |  |  |  |  |
| None | 141 (71.6%) | 86 (43.7%) | 3.6 | 1.9–7.0 | 0.001 |
| Primary | 35 (17.7%) | 64 (32.5%) | 0.8 | 0.5–2.2 | 0.012 |
| Secondary+* | 21 (10.7%) | 47 (23.8%) | Ref | – | – |
| **Father's Education** |  |  |  |  |  |
| None | 49 (24.9%) | 16 (8.1%) | 2.4 | 0.8–4.4 | 0.043 |
| Primary | 44 (22.3%) | 50 (25.4%) | 1.1 | 0.4–1.85 | 0.03 |
| Secondary+* | 104 (52.8%) | 131 (66.5%) | Ref | – | – |
| **Wealth Quintiles** |  |  |  |  |  |
| Poorest | 55 (27.9%) | 31 (15.7%) | 2.2 | 1.1–4.7 | 0.01 |
| Lower Middle | 49 (24.9%) | 28 (14.2%) | 2.5 | 1.2–5.4 | 0.03 |
| Middle | 34 (17.3%) | 45 (22.8%) | 1.1 | 0.5–2.3 | 0.01 |
| Upper Middle | 31 (15.7%) | 47 (23.9%) | 1.01 | 0.48–2.1 | 0.95 |
| Rich* | 28 (14.2%) | 46 (23.4%) | Ref | – | – |
| **Age of Child (Months)** |  |  |  |  |  |
| 6–11 | 8 (4.1%) | 12 (6.1%) | 1.1 | 0.4–3.3 | 0.86 |
| 12–23 | 90 (45.7%) | 51 (25.9%) | 5.1 | 2.3–11.2 | <0.001 |
| 24–35 | 62 (31.5%) | 52 (26.4%) | 3.7 | 1.7–8.4 | 0.001 |
| 36–47 | 24 (12.2%) | 44 (22.3%) | 1.8 | 0.7–4.6 | 0.20 |
| 48–59* | 13 (6.6%) | 38 (19.3%) | Ref | – | – |

\*Reference category

## DISCUSSION

Acute malnutrition is a key determinant of child health, impacting both physical and mental well-being [13]. This study explored the relationship between acute malnutrition and regional disparity, as well as socioeconomic factors. A significant association was found between regional disparity and acute malnutrition, with children in slum areas having 2.2 times higher odds of wasting compared to those in urban areas. This aligns with UNICEF’s National Nutritional Survey [3]. Slum areas, similar to rural regions in healthcare and education, are affected by factors like floods, climate change, and urban migration, which increase vulnerability to malnutrition. Younger children were more likely to experience malnutrition, possibly due to their higher dependency on maternal care [14]. While gender was not a significant factor in this study, males had 1.2 times higher odds of malnutrition, consistent with studies suggesting higher caloric needs in males [3,14]. Socioeconomic factors, including maternal and paternal education and household wealth, were strongly associated with malnutrition. Non-educated parents had higher odds of malnourished children, and children from the poorest households showed higher rates of malnutrition, confirming the role of economic constraints [15,16].

## Conclusion

The study concluded that acute malnutrition is linked to factors such as maternal and paternal education, area of residence, and household wealth. Addressing these modifiable factors, particularly by investing in girls’ education and promoting women’s empowerment, can help reduce malnutrition and improve child health.

Based on the study’s findings, further research is needed to explore determinants of acute malnutrition, particularly about regional disparities and socioeconomic determinants, as there is limited published data on this topic in Pakistan. Expanding the knowledge base will help identify root causes and inform policies to address underlying factors and regional variations in child health outcomes.

## Recommendations

Improving education, especially for mothers, should be prioritized. Collaboration with the government and local NGOs can help promote higher education. Additionally, enhancing healthcare in rural areas is crucial to reducing migration to urban centers. Together, these efforts will help reduce acute malnutrition, as discussed in the study.

Nutritional rehabilitation centers should be established nationwide in healthcare facilities to provide parents with regular health education and nutritional awareness. Public awareness campaigns should be implemented to advocate for better health practices among families.

A multi-sector approach, including public-private partnerships, is essential for improving infrastructure in slum areas, particularly in health and education.

## Acknowledgments

Dr. Shabana Junejo (AMS, CHK), Dr. Abdul Jabbar Memon (Principal, Public Health School Karachi).

## Availability of data

The data supporting the findings of this study are not publicly available due to institutional review board (IRB) and hospital policy restrictions regarding patient confidentiality.

